# CLINICAL AND MOLECULAR CHARACTERISATION OF SLC31A1-RELATED NEURODEVELOPMENTAL DISORDER

**DOI:** 10.1101/2024.11.27.24317634

**Authors:** Natalia Juliá-Palacios, Gerard Muñoz-Pujol, Reza Maroofian, Aida M. Bertoli-Avella, Marta Gómez-Chiari, Jordi Muchart, Abraham J. Paredes-Fuentes, Maria del Mar O’Callaghan, Irene S. Machado-Casas, Ingrid Cristian, Jennifer Morrison, Angels Garcia-Cazorla, Anna Codina, Mohammad Miryounesi, Emir Zonic, Peter Bauer, Huma Cheema, Muhammad Nadeem Anjum, Nouriya Al-Sannaa, Marwa Elmaksoud, Faroug Ababneh, Sahar Alijanpour, Seyed Hassan Tonekaboni, Afshin Fayazi, Maria Urbaniak, Uxia Barba, Janet Hoenicka, Francesc Palau, Henry Houlden, Juan Darío Ortigoza-Escobar, Antonia Ribes, Carlos Santos-Ocaña, Millie Tyler, Patrick Gaffney, Christopher Carroll, Frederic Tort, Klaas J. Wierenga, Bryn Webb, Rafael Artuch, Heidy Baide-Mairena, Roser Urreizti

## Abstract

Copper is indispensable for various metabolic processes, notably mitochondrial respiration. In humans, copper homeostasis hinges on transporters such as copper transporter 1 (CTR1), encoded by the *SLC31A1* gene. Recently, bi-allelic mutations in *SLC31A1* have been associated with a new neurodevelopmental disorder. This study presents clinical, genetic, and biochemical findings from 13 new cases across 10 families worldwide. RNA sequencing evaluated gene expression, and Western blotting assessed CTR1 protein levels. Additionally, mitochondrial respiratory capacity was measured via high-resolution respirometry. Affected individuals exhibited a distinct clinical phenotype characterized by early-onset epileptic encephalopathy, severe neurodevelopmental delay and hypotonia, with high mortality. Neuroimaging revealed significant brain atrophy and white matter abnormalities. Genetic analysis identified bi-allelic *SLC31A1* variants, predominantly p.His120Gln in six cases and p.(Arg102Cys/His) in three cases. Functional studies in patient fibroblasts demonstrated impaired mitochondrial respiration. This study significantly broadens the clinical spectrum of this recently described syndrome, presenting as a severe developmental encephalopathy with high mortality risk, and suggests mitochondrial dysfunction as a potential pathomechanism. These findings contribute to the mounting evidence linking CTR1 dysfunction to neurodegeneration, underscoring the urgency for further therapeutic investigations.

## INTRODUCTION

Copper (Cu) is an essential metal that is required for the proper functioning of metabolic pathways, including the synthesis of collagen and catecholamines. It is also the cofactor of cytochrome c oxidase in the mitochondrial respiratory chain, and of the antioxidant enzyme superoxide dismutase, among others. Thus, copper plays an important role for cellular respiration, antioxidant defense, connective tissue formation, neurotransmitter biosynthesis, neuropeptide amidation and iron homeostasis.^1, 2^ There is evidence of copper homeostasis contribution in developmental and reparative angiogenesis.^3^ Intracellular Cu concentration is tightly regulated by dedicated proteins that facilitate its uptake, efflux, and distribution to target Cu-dependent proteins and enzymes.^4^ Two families of membrane proteins are critical for Cu homeostasis in mammals: the P-type ATPase copper pumps, ATP7A and ATP7B (MIM *300011 and *606882), that move Cu against a concentration gradient during Cu secretion or intracellular sequestration, and copper transporters (CTR) that are responsible for the initial uptake of Cu into cells. *AP1S1* (MIM *603531), encodes the small subunit σ1A of the adaptor protein-1 (AP1) complex, necessary for the intracellular trafficking of ATP7A. Through its involvement in clathrin-coated vesicle assembly it directs the intracellular trafficking of copper pumps ATP7A and ATP7B.^5^ In humans, two CTR transporters have been identified, CTR1 (O15431) and CTR2 (O15432), encoded by the *SLC31A1* and *SLC31A2* genes, respectively (MIM *603085 and *603088). CTR1 is 190 amino acids long and forms a homotrimeric transmembrane transporter that is ubiquitously expressed, with relatively high levels in liver, small intestine, and brain (Lee *et al*^6^ and GTEx Portal accessed August 2024). CTR1 is an endothelial Cu importer and functions as a redox sensor to promote angiogenesis in endothelial cells as it has been demonstrated that oxidation of CTR1 at Cys189 promotes vascular endothelial growth factor receptor type 2 (VEGFR2) internalization and signaling to enhance angiogenesis in mice.^3^

Hitherto, different genetic disorders have been reported regarding Cu homeostasis. These include well-known Menkes disease (MIM #309400) and Wilson disease (MIM #277900), caused by pathogenic variants at *ATP7A* and *ATP7B* genes respectively, aceruloplasminemia (MIM #604290) due to mutations in *CP* (MIM *117700), MEDNIK syndrome (MIM # 609313), a multisystemic disease that combines clinical and biochemical signs of both Menkes and Wilson’s diseases related to genetic defects in *AP1S1*, MEDNIK-like syndrome (MIM #242150) caused by mutations in *AP1B1* (MIM *600157),^7^ Huppke-Brendel syndrome (MIM #614482) associated with a severe loss of function of the endoplasmic reticulum (ER) membrane acetyl-CoA transporter 1 (AT-1) protein, encoded by *SLC33A1* (MIM *603690),^8^ and the very recently identified NSCT (Neurodegeneration and seizures due to copper transport defect, MIM #620306) caused by recessive mutations in *SLC31A1*. Pathogenic variants in this gene have been reported in three cases with a deficiency in the high-affinity copper uptake protein 1 (COPT1; CTR1; O15431).^9, 10^ This newly described genetic disorder leads to neurodegeneration and seizures and presents in the neonatal period or early infancy. Evidence of impaired intracellular Cu homeostasis was reported with intense expression of CTR1 in the basolateral aspect of the polarized choroid plexus epithelium, which mediates Cu uptake in the brain.^9^

Here we present thirteen new cases from ten different families. Among them, five cases from four unrelated families are homozygous for the same novel pathogenic variant in *SLC31A1* (NM_001859.4:c.360C>G; p.His120Gln). Through our comprehensive analysis, we describe the phenotype of the SLC31A1 deficiency associated syndrome. Additionally, biochemical and molecular studies were conducted to further elucidate the underlying mechanisms of this condition.

## RESULTS

### Clinical Findings

The clinical features, biochemical and genetic data from thirteen cases (7 males / 6 females) are summarized in Table 1. Further description of the clinical presentation of each subject (and HPO descriptions) is included as Supplementary Clinical Information and Supplementary Table 1.

**Table 1.**
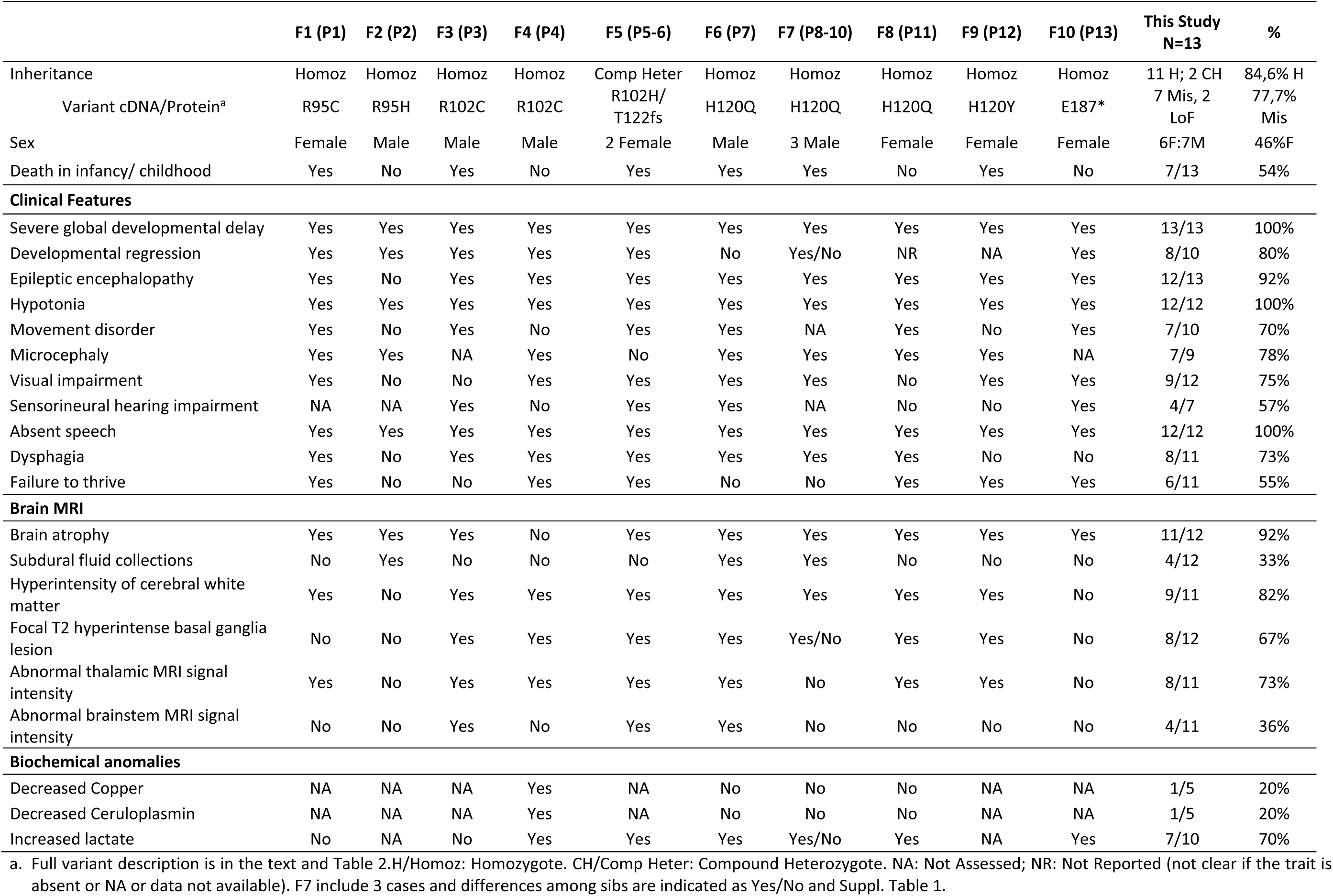
Main clinical characteristics of the thirteen cases described here.

|  | F1 (P1) | F2 (P2) | F3 (P3) | F4 (P4) | F5 (P5-6) | F6 (P7) | F7 (P8-10) | F8 (P11) | F9 (P12) | F10 (P13) | This Study<br>N=13 | % |
| --- | --- | --- | --- | --- | --- | --- | --- | --- | --- | --- | --- | --- |
| Inheritance | Homoz | Homoz | Homoz | Homoz | Comp Heter | Homoz | Homoz | Homoz | Homoz | Homoz | 11 H; 2 CH | 84,6% H |
| Variant cDNA/Protein<sup>a</sup> | R95C | R95H | R102C | R102C | R102H/<br>T122fs | H120Q | H120Q | H120Q | H120Y | E187* | 7 Mis, 2<br>LoF | 77,7%<br>Mis |
| Sex | Female | Male | Male | Male | 2 Female | Male | 3 Male | Female | Female | Female | 6F:7M | 46%F |
| Death in infancy/ childhood | Yes | No | Yes | No | Yes | Yes | Yes | No | Yes | No | 7/13 | 54% |
| **Clinical Features** |  |  |  |  |  |  |  |  |  |  |  |  |
| Severe global developmental delay | Yes | Yes | Yes | Yes | Yes | Yes | Yes | Yes | Yes | Yes | 13/13 | 100% |
| Developmental regression | Yes | Yes | Yes | Yes | Yes | No | Yes/No | NR | NA | Yes | 8/10 | 80% |
| Epileptic encephalopathy | Yes | No | Yes | Yes | Yes | Yes | Yes | Yes | Yes | Yes | 12/13 | 92% |
| Hypotonia | Yes | Yes | Yes | Yes | Yes | Yes | Yes | Yes | Yes | Yes | 12/12 | 100% |
| Movement disorder | Yes | No | Yes | No | Yes | Yes | NA | Yes | No | Yes | 7/10 | 70% |
| Microcephaly | Yes | Yes | NA | Yes | No | Yes | Yes | Yes | Yes | NA | 7/9 | 78% |
| Visual impairment | Yes | No | No | Yes | Yes | Yes | Yes | No | Yes | Yes | 9/12 | 75% |
| Sensorineural hearing impairment | NA | NA | Yes | No | Yes | Yes | NA | No | No | Yes | 4/7 | 57% |
| Absent speech | Yes | Yes | Yes | Yes | Yes | Yes | Yes | Yes | Yes | Yes | 12/12 | 100% |
| Dysphagia | Yes | No | Yes | Yes | Yes | Yes | Yes | Yes | No | No | 8/11 | 73% |
| Failure to thrive | Yes | No | No | Yes | Yes | No | No | Yes | Yes | Yes | 6/11 | 55% |
| **Brain MRI** |  |  |  |  |  |  |  |  |  |  |  |  |
| Brain atrophy | Yes | Yes | Yes | No | Yes | Yes | Yes | Yes | Yes | Yes | 11/12 | 92% |
| Subdural fluid collections | No | Yes | No | No | No | Yes | Yes | No | No | No | 4/12 | 33% |
| Hyperintensity of cerebral white matter | Yes | No | Yes | Yes | Yes | Yes | Yes | Yes | Yes | No | 9/11 | 82% |
| Focal T2 hyperintense basal ganglia lesion | No | No | Yes | Yes | Yes | Yes | Yes/No | Yes | Yes | No | 8/12 | 67% |
| Abnormal thalamic MRI signal intensity | Yes | No | Yes | Yes | Yes | Yes | No | Yes | Yes | No | 8/11 | 73% |
| Abnormal brainstem MRI signal intensity | No | No | Yes | No | Yes | Yes | No | No | No | No | 4/11 | 36% |
| **Biochemical anomalies** |  |  |  |  |  |  |  |  |  |  |  |  |
| Decreased Copper | NA | NA | NA | Yes | NA | No | No | No | NA | NA | 1/5 | 20% |
| Decreased Ceruloplasmin | NA | NA | NA | Yes | NA | No | No | No | NA | NA | 1/5 | 20% |
| Increased lactate | No | NA | No | Yes | Yes | Yes | Yes/No | Yes | NA | Yes | 7/10 | 70% |

a. Full variant description is in the text and Table 2. H/Homoz: Homozygote. CH/Comp Heter: Compound Heterozygote. NA: Not Assessed; NR: Not Reported (not clear if the trait is absent or NA or data not available). F7 include 3 cases and differences among sibs are indicated as Yes/No and Suppl. Table 1.

All patients presented with severe neurological impairment characterized by early onset epileptic encephalopathy (EOEE), with an average age of onset of 6 months (range 1–24 months). Intractable polymorphic seizures, described as myoclonic, tonic, clonic, and infantile spasms, were observed. Only three out of 13 patients had normal neurodevelopment prior to seizure onset. Physical examination revealed severe hypotonia without head control in all 13 cases, along with microcephaly in seven out of nine. Additionally, movement disorders (tremor, parkinsonism, myoclonus, generalized chorea, and generalized dystonia) were present in seven of ten cases. Visual impairment (blindness, optic disc pallor, abnormal ocular movements, strabismus, cataracts) was noted in nine out of twelve cases, and sensorineural hearing loss in four out of seven. As the disease progressed, most developed dysphagia (8/11), and one required ventilatory support. Seven cases died prematurely at an average age of 2.7 years (range 1–6 years).

Metabolic investigations revealed elevated lactate levels in plasma (n=6/10) and CSF (n=5/5) with increased pyruvate and alanine in one case. Serum copper and ceruloplasmin were normal in the four tested individuals, although one showed decreased circulating ceruloplasmin concentration.

Twelve patients received brain MRI, including three with follow-up imaging (Fig.1, Table 1, and Supplementary Table 1). All affected individuals but one showed global brain atrophy, and compensatory enlargement of the ventricles in nine. Thinning of the corpus callosum or enlarged subarachnoid spaces were seen in six. Other common features included T2-WI hyperintensity involving the white matter, thalamus, basal ganglia, or brainstem. Radiological findings suggesting vigabatrin toxicity were observed in two cases. Intracranial hemorrhage was present in four individuals, involving the subarachnoid space, basal ganglia, or in the form of subdural hematomas. Other subdural fluid collections, such as hygromas, were observed in three cases.

**Figure 1:**
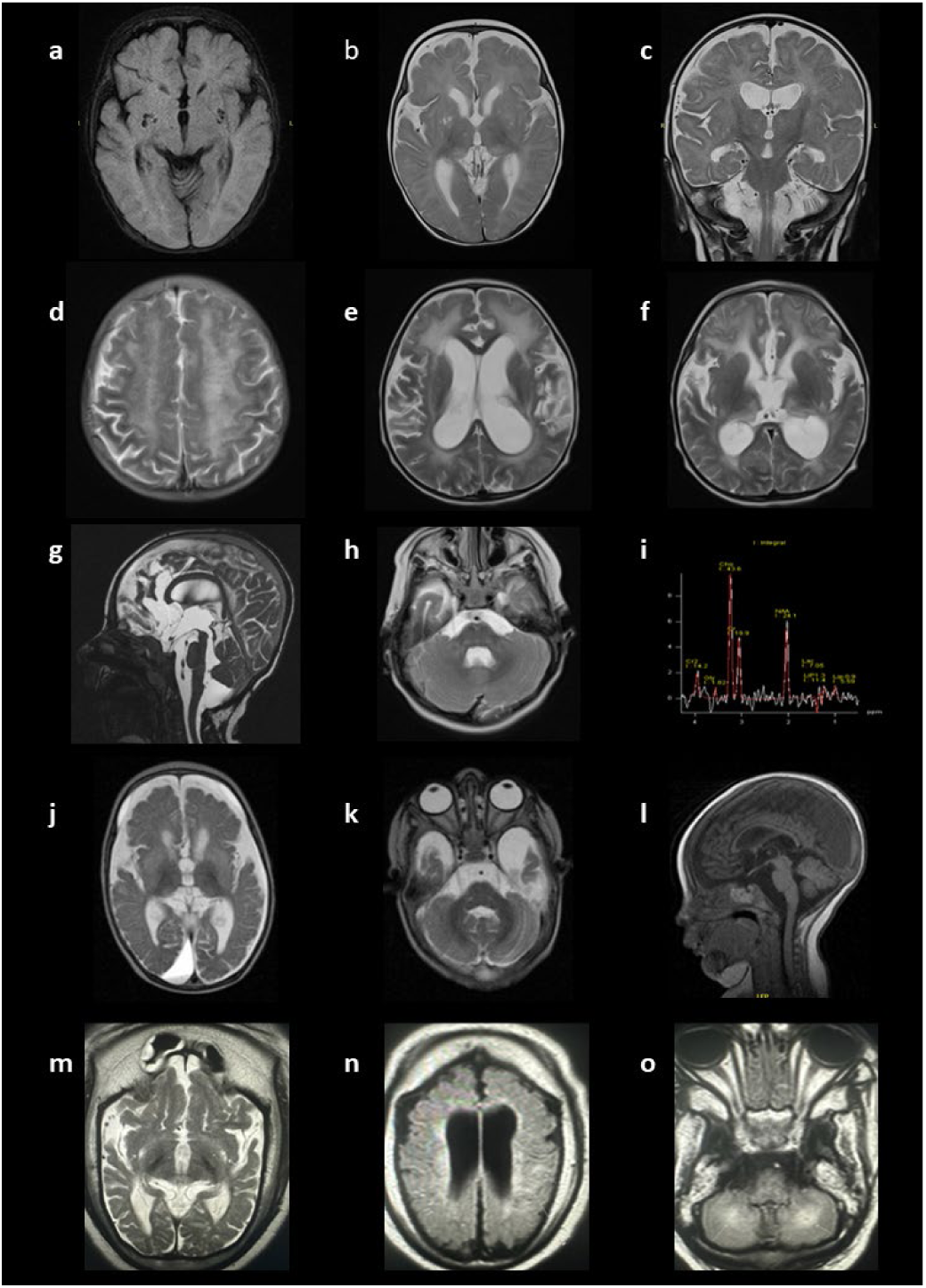
Brain MRIs of cases P1 (a to c), P3 (d to i), P7 (j to l) and P9 (m to o). **P1** at 3-6 months of age **(a)** Prominent Virchow-Robin spaces can be seen in the lenticular nuclei and subganglionar area (axial, FLAIR T2). **(b)** Enlargement of the ventricular system and extraxial spaces without subdural fluid collections and corpus callosum thinning (axial, FSE T2) **(c)** High T2 signal intensity in the pulvinar nuclei of the thalami (coronal, FSE T2). Myelination is delayed for a 3 to 6-months-old boy. **P3** at 2-4 years of age: **(d)** Symmetrical hyperintensities in bilateral centrum semiovale with a tigroid pattern (axial, T2-weighted); **(e)** Ventriculomegaly, hyperintensity in white matter periventricular and external capsule (T2-weighted axial), **(f)** Loss of volume and abnormal signal in the thalamus (T2-weighted axial), **(g)** Ventriculomegaly without obstruction, and megacisterna magna (CISS sagittal), **(h)** Abnormal signal in the posterior pons and bilateral middle cerebellar peduncle (T2-weighted axial) and **(i)** Decrease in the NAA peak observed in the MR spectroscopy (voxel placed in deep white matter with an echo time of 144 ms). **P7** at 6-9 months of age: **(j)** Hyperintense signal abnormalities in both the basal ganglia and diffuse brain atrophy prominently at bilateral frontotemporal lobes with secondary ventricular dilatation and frontal bilateral and right occipital subdural hygroma (T2-weighted axial) **(k)** Hyperintense signal abnormalities are observed in the bilateral brainstem, including severe temporal cortical-subcortical atrophy with significant enlargement of the subarachnoid space of the anterior temporal fossa. **(l)** Compensatory enlargement of the ventricular system and thinning of the corpus callosum (T1-weighted sagittal images). **P9** at 6-9 months of age **(m, n)** Severe frontotemporal cortical-subcortical atrophy with significant enlargement of the subarachnoid space of the bihemispheric convexity associated with frontoparietal subdural hygromas along with compensatory enlargement of the ventricular system and prominent thinning of the corpus callosum, **(o)** T2-WI hyperintense signal was observed throughout the upper left parietal lobe and medial cerebellar hemispheres (white arrows).

### Genetic findings

The thirteen cases presented here were found to have bi-allelic disease-causing variants in *SLC31A1*. We have identified eight different variants, of which seven were novel (Tables 1 and 2). Nomenclature for each variant is determined using the canonical *SLC31A1* transcript NM_001859.4/ ENST00000374212. Ten cases are homozygotes for missense variants p.Arg95His, p.(Arg95Cys), p.(Arg102Cys), p.His120Gln and p.(His120Tyr), one is homozygote for the premature stop codon p.(Glu187*) and two siblings are compound heterozygotes for the c.363_364dupAA; p.(Thr122LysfsTer8) and c.304C>T; p.(Arg102His) variants. The genetic position, in silico prediction scores, and ACMG classification of the variants reported here, along with previously published variants, are summarized in Table 2.

**Table 2.**
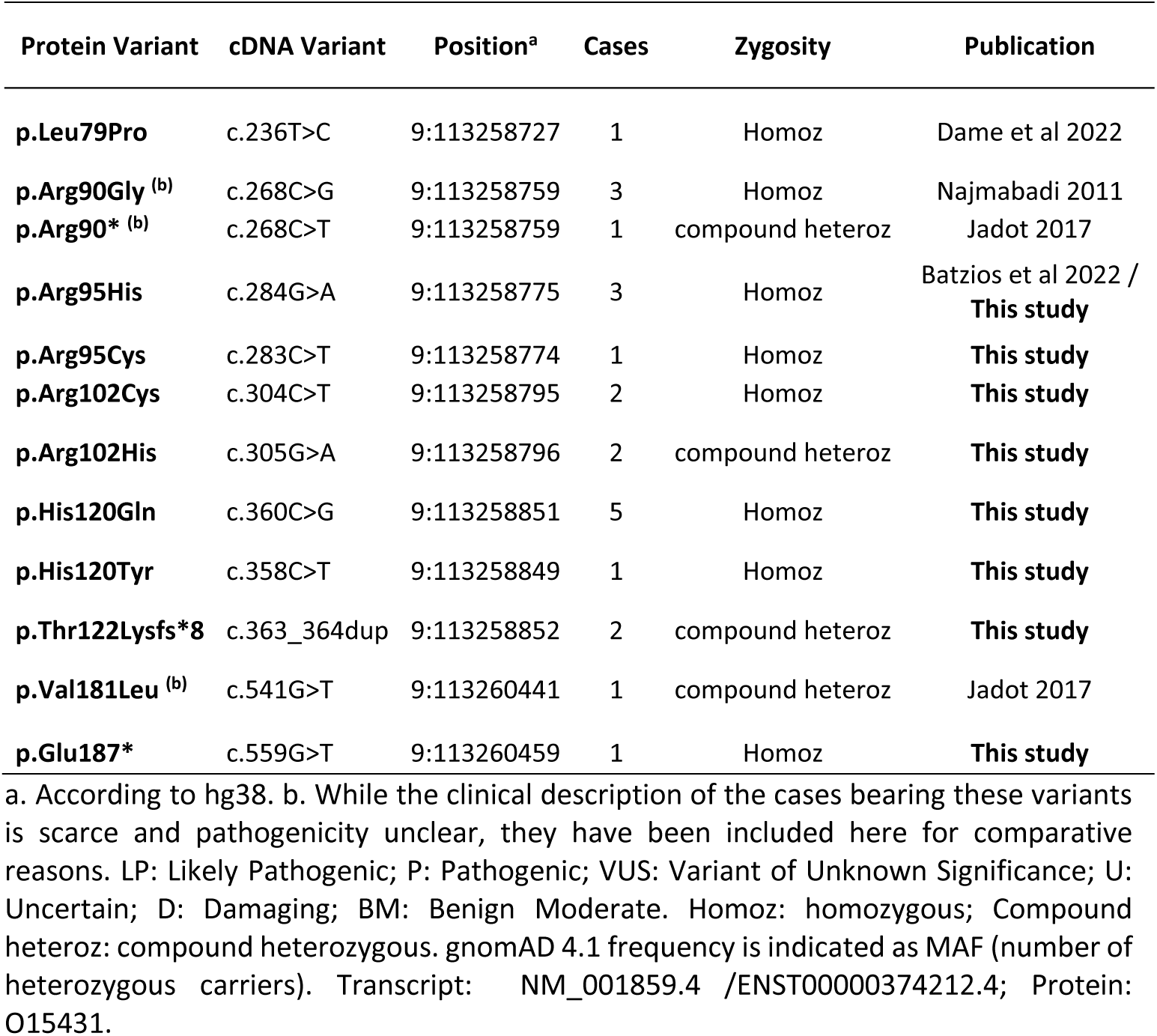
*SLC31A1* pathogenic variants described here and previously published.

All the missense variants affect highly conserved residues, conserved in all the assessed vertebrates (Supplementary Fig. 1), are absent or at a very low frequency in gnomAD (v. 4.1, accessed August 2024). We evaluated the gene-disease relationship (GDR) evidence following the Clinical Genome Resource (ClinGen) Clinical Validity Framework.^11^ Briefly, the strength of evidence for a GDR can be categorized as Definitive, Strong, Moderate, Limited, or No Known Disease Relationship. This is relevant because only genes with evidence of Moderate or above should be included in clinical diagnostic testing (add ref. PMID: 31732716). For *SLC31A1* we determined a Moderate level of evidence, confirming its association to autosomal recessive *SLC31A1*-related neurodevelopmental disorder.

Variant c.283C>T (p.Arg95Cys), identified in **P1**, affects the same residue as the previously reported p.Arg95His also identified here in **P2**.^9^ This variant affects an alpha-helix in the cytoplasmic gate of the CTR1 channel ( Fig. 2), close to the c.304C>T; p.(Arg102Cys) variant, identified here in two cases (**P3** and **P4**), while **P5** and **P6** are confirmed compound heterozygotes for the p.(Arg102His) variant that affects the same residue and the c.363_364dupAA variant that is predicted to skip the NMD process and be translated into the C-terminus truncated protein p.(Thr122Lysfs*8). Five cases from three unrelated families (**P7-11**) bear the same transversion c.360C>G, which leads to a missense change of the electrically charged His to the non-charged hydrophilic Gln at residue 120 (p.His120Gln). The three families are of Hispanic origin (Spain and Mexico), suggesting a potential founder effect. **P12** bears the c.358C>T change, affecting the same residue, p.(His120Tyr). His120 lies in the cytoplasmic region of the protein, in close vicinity to the CHC domain at the C-terminal end of the protein (Fig. 2). Finally, **P13** has the c.559G>T variant, which results in predicted protein truncation at p.(Glu187*). This variant leads to the loss of the last three residues in the 190-residue protein, the CHC domain. Case **P4**, an individual of Egyptian origin, is homozygous for a missense variant in the *ATP7B* gene, which encodes a copper-transporting ATPase associated with Wilson disease (WD; MIM #277900). The variant identified, c.1646T>C; p.(Leu549Pro), has been previously reported in another Egyptian patient with WD^12^ and is classified as VUS according to ACMG criteria ( PM2, PP3). As functional studies were not conducted on this variant by Abdelghaffar et al.,^12^ and this region of ATP7B does not show significant constraint against missense variants (Z = -0.33, regional constraint according to gnomAD 4.1), its pathogenicity remains unclear and the potential interaction between both etiologies should be considered in the case of **P4**.

**Figure 2:**
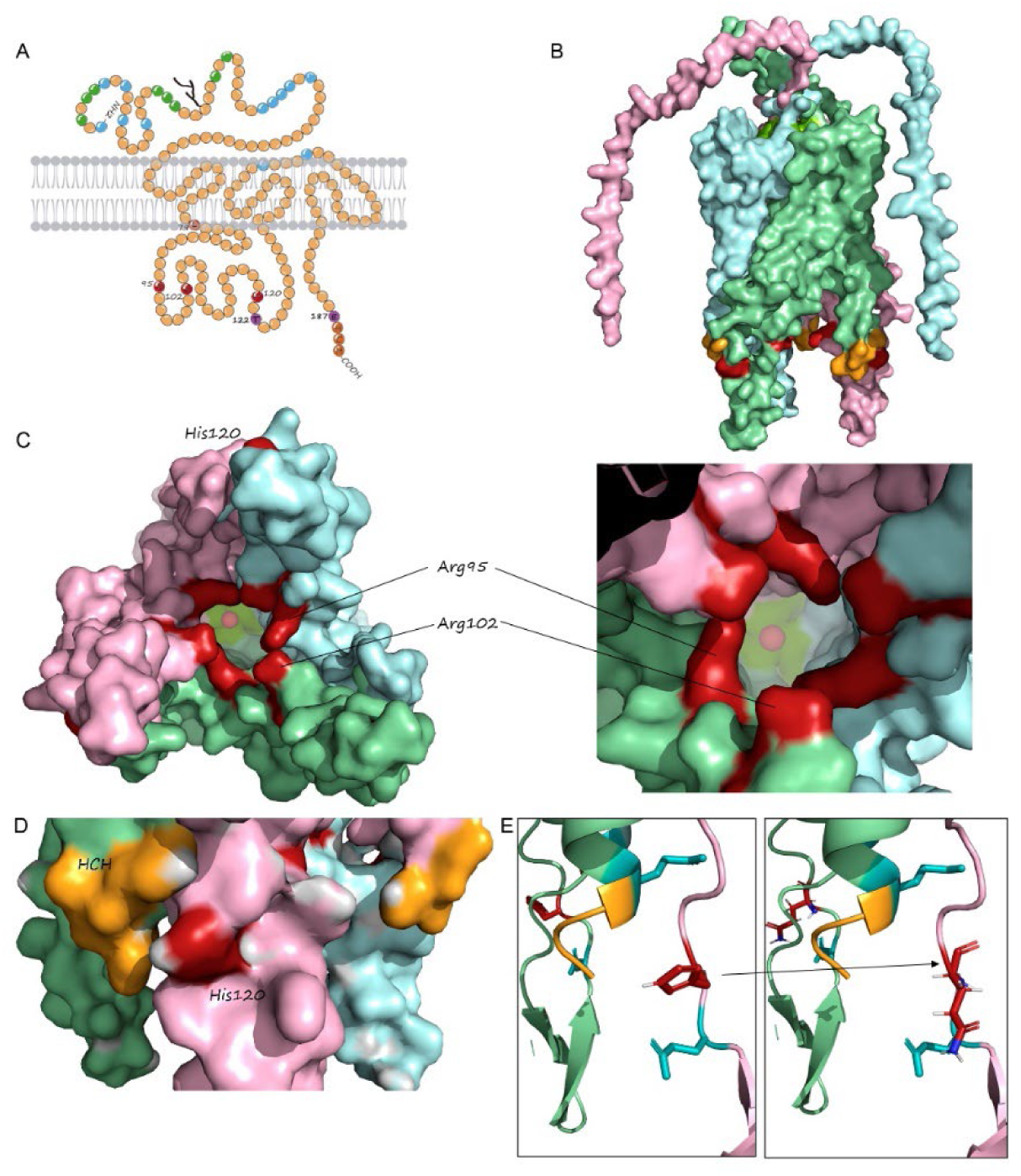
CTR1 protein. **A)** Topological model of the human CTR1 protein. His and Met-rich domains are indicated in green and blue, respectively, and the HCH C-term domain is shown in orange. The residues Leu79, Arg95, Arg102, and His120, mutated in the SLC31A1 cases, are coloured red and position of truncating variants at Thr122 and Glu187 are in violet. **B)** Lateral view of the surface of the trimeric hCTR1, each monomer in a different color. Mutated residues are shown in red, while the HCH domain is shown in orange. **C)** Bottom (intracellular) view of the trimer showing Arg 95 and 102 at the “gate” of the pore and His120 at the external surface. Copper is shown inside the pores as a red sphere. **D)** Detailed lateral view showing His120 in close vicinity to HCH domain. **E)** Modeled substitution of His120 by Gln.

### Functional studies

RNA-seq was performed on **P7** fibroblasts and the *SLC31A1* expression levels were not significantly altered in comparison with the entire cohort of transcriptomes (Supplementary Fig. 2). Expression of genes related to Cu metabolism or transport that have been altered in other Cu-related pathologies was also within the normal range, including *ATOX1* and *CCS*, the cytoplasmic chaperones involved in Cu reception (Supplementary Fig. 3A and 3B). Also, total CTR1 protein was assessed by Western blot in fibroblasts (**P7**) and lymphocytes (**P9**), both bearing the homozygous variant c.360C>G (p.His120Gln), and no differences were observed in these cell lines when compared with controls (Suppl. Fig. 5). Cu intracellular uptake was analyzed in fibroblasts of **P7** and no significant alterations were observed compared with controls studied in parallel (data not shown).

The mitochondrial respiratory capacity of **P7** fibroblasts was investigated by analyzing the oxygen consumption rate (OCR) by high-resolution respirometry. Results showed an important impairment in **P7**’s fibroblasts basal respiratory rate compared to control cells (*p*<0.05). In addition, **P7** cells also had significant reduction in their maximal respiratory capacity (*p*<0.05) when treated with the mitochondrial uncoupler CCCP (Fig. 3). Treatment of **P7** cells with 50 μM copper histidinate (CuHis) for 24 h did not show any significant respiration improvement. However, increasing treatment dose up to 100 μM CuHis for 72 h resulted in a slight but significant improvement in respiratory capacity for the basal respiratory rate (*p*<0.05) and for the maximal respiratory capacity (*p*<0.05) (Fig. 3).

**Figure 3:**
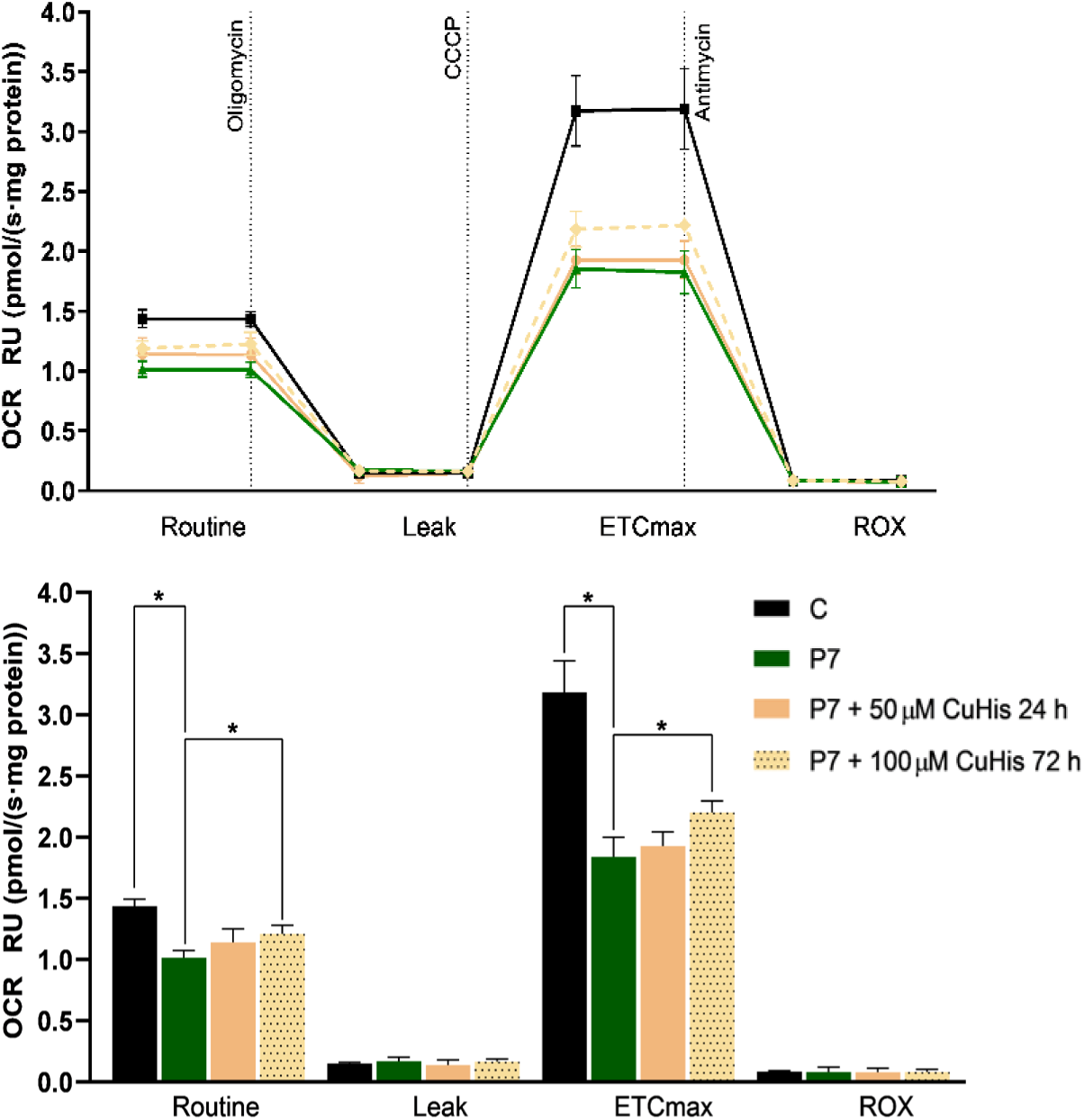
Mitochondrial function assessment. Mitochondrial respiration is reduced in P7 fibroblasts. Mitochondrial respiratory capacity was analyzed in P7 fibroblasts by investigating the oxygen consumption rate (OCR) with high-resolution respirometry. The upper panel shows a scatter plot of mean OCR results across four time intervals delimited by the injection of the three compounds oligomycin, CCCP (carbonyl cyanide 3-chlorophenylhydrazone), and antimycin A. Dotted lines indicate the exposure of cells to each compound. The lower panel represents the mean results ± SD as a bar chart with statistical results. A significant decrease in basal OCR was observed in P7 cells compared to the control. P7 fibroblasts also exhibited a significant reduction in maximal respiratory capacity (*p*<0.05) upon CCCP treatment. Treatment with 100 μM CuHis for 72 h, but not with 50 μM CuHis for 24 h, partially restored the mitochondrial respiratory capacity in P7 fibroblasts. Both basal and ETCmax respiratory rates were significantly increased (*p*<0.05) upon 100 μM CuHis treatment (72 h) compared to non-treated P7 cells. C: control; CuHis, copper histidinate. OCR at basal state (ROUTINE); residual oxygen consumption after oligomycin treatment (LEAK); maximum oxygen consumption induced by CCCP titration (ETCmax); residual oxygen consumption after antimycin A treatment (ROX). OCR was measured as pmol/(second * mg of protein). Data are expressed as relative units (RU) of non-treated P7 cells. Two biological replicates were performed for each experiment.

## DISCUSSION

Here we present the clinical and molecular findings of thirteen affected children with a recently described developmental encephalopathy caused by bi-allelic *SLC31A1* variants. The same homozygous variant (c.360C>G, p.His120Gln) was identified in three families. In most cases, the disease presented in the first year of life as a severe early onset epileptic encephalopathy (EOEE) resulting in profound disability leading to early death before three years of age in half of the cases, supporting the high mortality risk associated with this clinical entity. Seven cases with recessive *SLC31A1* pathogenic variants have been reported in the literature.^9, 10, 13, 14^ Comprehensive descriptions of the *SLC31A1*-related syndrome have been published in three individuals (Table 3)^9, 10^ while phenotypic traits for four of them are not available.^13, 14^

**Table 3.**
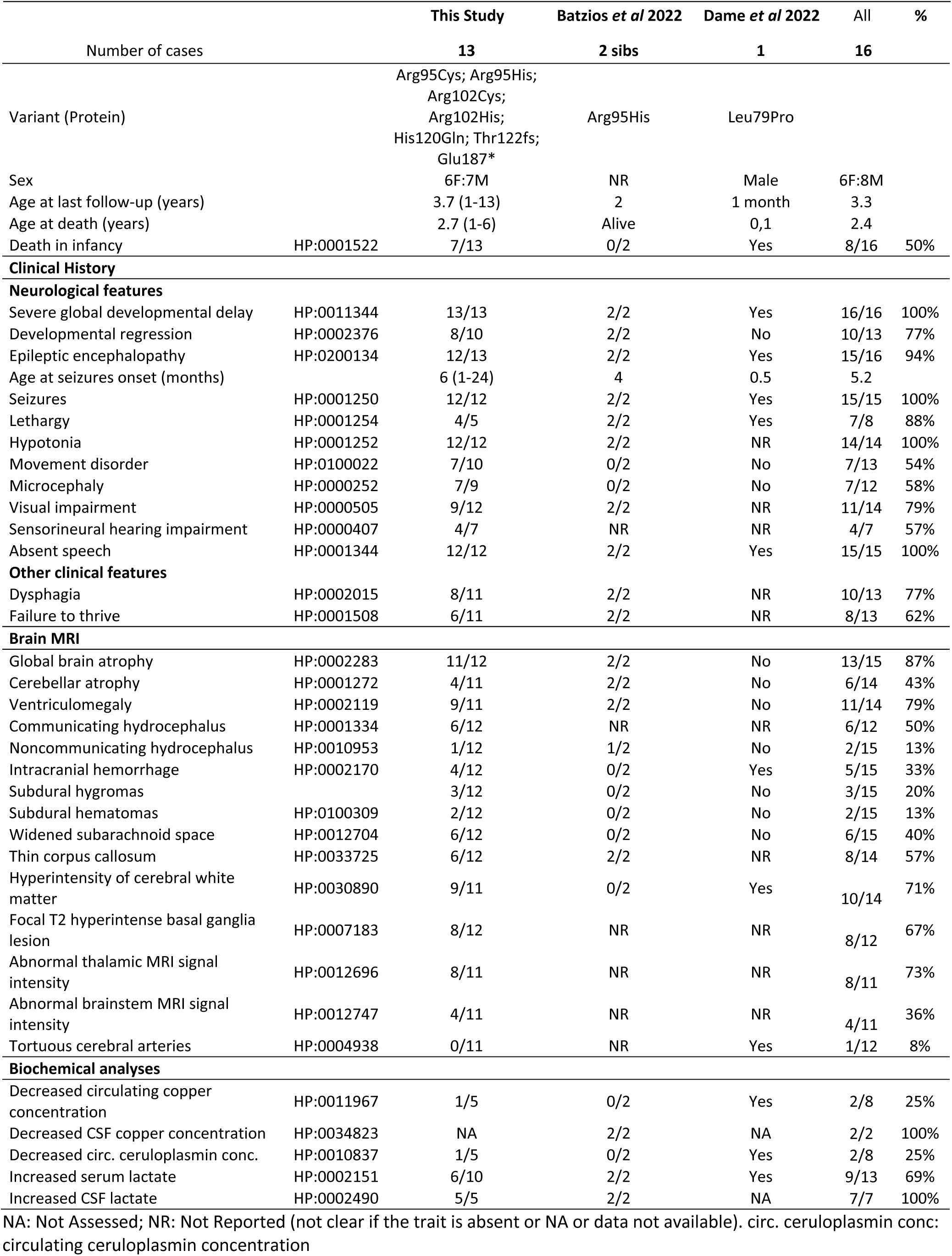
Overview of the main characteristics of the 16 cases described so far.

The clinical presentation in the cases reported here closely resembles that of the twins documented by Batzios *et al*.^9^ (Table 3), who were homozygotes for the p.Arg95His variant, but contrasts with the congenital presentation observed in the case described by Dame et al.,^10^ who was homozygote for the p.(Leu79Pro) variant. The latter presented with major congenital anomalies involving the pulmonary, vascular, skeletal, and central nervous systems, seizures at two weeks of life, and acute intracranial hemorrhages diagnosed with cranial ultrasound. This proband appears to have the most severe presentation of this disease to date, and is the only proband known to have low levels of serum copper and ceruloplasmin. Due to the retrospective nature of this report and the recent identification of this disorder, copper and ceruloplasmin levels were not assessed in most cases. However, these levels were measured in four out of five individuals harboring the c.360C>G (p.His120Gln) variant and all showed normal values. Case **P4**, who bears the p.(Arg102Cys) *SLC31A1* variant in addition to the p.(Leu549Pro) *ATP7B* variant, was identified to have low levels of serum copper and ceruloplasmin, together with low urine copper levels, the latter finding contrary to what would be expected in WD. Urine copper in other cases were not assessed so it is unknown if SLC31A1 deficiency is responsible for his low copper urine excretion and to which extent these two variants are altering the copper transport in this patient. **P12** is homozygote for the NM_000018.4:c.1246G>A p.(Ala416Thr) variant in the very long-chain acyl-CoA dehydrogenase gene (ACADVL *609575), which is associated with VLCAD deficiency (ACADVLD; MIM # 201475) (see supplementary clinical description). This particular variant has been previously described as associated with mild or late-onset (adult form) disease, and functional analysis revealed that this variant maintains moderate activity.^15–18^ While this likely pathogenic variant may be modifying this proband’s phenotype to some extent, it does not seem responsible for the main clinical presentation. These two cases highlight the complexity in understanding and managing clinical presentations with dual diagnoses. Further functional studies would be necessary to fully understand the implication of each disease gene involved.

Biomarkers indicative of mitochondrial impairment were observed not only in our cohort but also in previously reported patients, reflecting the well-established role of copper (Cu) in mitochondrial metabolism. Notably, serum lactate was elevated in 60% of the analyzed cases, and cerebrospinal fluid (CSF) lactate was elevated in all analyzed cases, consistent with findings in previously reported cases.^9, 10^ This mitochondrial dysfunction appears to significantly impact the clinical outcomes of the patients. Moreover, impaired mitochondrial respiratory chain activities in muscle and impaired mitochondrial respiration in fibroblasts were detected in the patient reported by Batzios et al.^9^ and in **P7**, increased alanine in blood and CSF, elevated serum GDF-15 and FGF-21 and Krebs cycle metabolites in urine were detected suggesting mitochondrial disease and leading to cofactors supplementation in this patient. Additionally, Leigh syndrome was suspected in two individuals who experienced developmental regression following infantile spasms, elevated lactate levels in both plasma and cerebrospinal fluid (CSF), and symmetrical T2-weighted hyperintensities in the basal ganglia, thalamus, and dentate nuclei. Therefore, defects in copper transporter receptors due to pathogenic variants in *SLC31A1* should be considered in the differential diagnosis of mitochondrial diseases.

The cases presented here display similarities with the clinical spectrum observed in other intracellular copper trafficking disorders, including rapid progressive neurodegeneration, intractable seizures, hypotonia, microcephaly, and severe brain atrophy since early infancy. These findings were frequently described in patients with Menkes disease,^19^ and Huppke-Brendel Syndrome.^8^ Additionally, white matter hyperintensities in the bilateral centrum semiovale and subdural collections, such as hygromas and/or hematomas, are also described in Menkes disease.^19^ However, the subjects reported here did not show other typical neurocutaneous signs including brittle hair, hypopigmentation, and connective tissue laxity as seen in Menkes, nor the ichthyosis and keratoderma observed in MEDNIK syndrome.^5^ Furthermore, there were no signs of tissue copper overload, such as hepatic cirrhosis, liver failure, or ocular Kayser-Fleisher rings, which are characteristic of Wilson disease.^20–23^

To date, experiments conducted on fibroblasts from patients with *SLC31A1* mutations (Batzios et al^9^ and this study) have not revealed significant disruptions in CTR1 protein expression or localization. This suggests that, for most pathogenic variants, copper (Cu) impairment is likely not primarily due to cellular import. Instead, it is probably related to its delivery and trafficking within the cell, particularly to the mitochondrion (note that only total intracellular copper, not intramitochondrial copper, was measured). Alternatively, other mechanisms not directly involving copper, where CTR1 cysteine residues play a key role, may be involved.^3^ Our study does not show an improvement in mitochondrial respiratory capacity in response to treatment with copper histidinate, unlike what was previously observed.^9^ It is unclear if this lack of response is due to methodological issues or to the particular variant and its effect on the CTR1 protein function. In this sense, it is interesting to notice the different positions of these mutations in the CTR1 channel. The CTR1 active channel is a transmembrane homotrimer (Fig. 2) that forms a pore that will import Cu from the extracellular space to the cytoplasm with high affinity.^24^ It has been proposed that four methionine residues (43, 45, 150 and 154) are involved in Cu transport through the core, and the cytoplasmic domain is involved in the controlled delivery of Cu to the corresponding chaperones (ATOX1, CCS, MT, etc.), which will distribute Cu intracellularly.^25^ All the variants identified in this study affect the first intracellular domain of CTR1 (residues 83-132; Fig. 1). Residue Arg95 lies at the “gate” of the internal cavity of the CTR1 pore (Fig. 1),^26^ and its mutation for His has been proposed to alter the internal charges and Cu affinity.^9^ This residue is stabilized by H bonds with Arg91, and its substitution by His or Cys (like in the patients presented here) could impact both the intrapore electric charge and stability as well as the gate integrity. Previous studies showed that the substitution of His139 by Arg increased CTR1 *Km* values, demonstrating the relevance of the Arg and His equilibrium in the CTR1 Cu transport.^27, 28^ The Arg102 also lies in the cytoplasmic domain, in close vicinity to Arg95 in the tridimensional CTR1 model (Fig. 2). Again, the substitution of the Arg by His or Cys seems to be disturbing the Cu transport through the pore and its delivery to the appropriate chaperones. His120 is also on the cytoplasmic loop of CTR1, but far from the gate. The 3D protein model locates this residue in close proximity to the HCH domain of the neighboring monomer. This domain has been proven to be determinant in the delivery of Cu to the intracellular chaperones, especially ATOX1.^25^ In contrast, p.(Leu79Pro), described in the only patient presenting with low copper and ceruloplasmin circulating levels, introduces a proline residue in an alpha-helix of the first transmembrane domain, a change highly structurally disturbing, and probably affecting the pore shape and stability and thus completely blocking Cu import (Fig. 1). The only patients bearing an early truncating variant (P5 and P6 sisters) were also bearing the p.(Arg102His) variant and ceruloplasmin levels were not assessed (both passed away and further functional studies are not available). To date, only 16 patients have been described, and clinical information remains limited, particularly regarding circulating copper and ceruloplasmin levels. Nevertheless, a genotype-phenotype correlation is emerging. Severe variants that disrupt the pore structure of the CTR1 channel appear to be associated with a more severe phenotype, characterized by low circulating copper and ceruloplasmin levels. In contrast, cytoplasmic mutations, potentially impacting intracellular copper trafficking, seem to result in a milder phenotype with copper and ceruloplasmin levels mostly within normal ranges. However, both forms are associated with mitochondrial dysfunction, epileptic encephalopathy, hypotonia, severe brain atrophy and death during early childhood.

In summary, the comprehensive characterization of 13 patients with recessive *SLC31A1* variants, including the recurrent c.360C>G (p.His120Gln) variant, strengthens the association between CTR1 dysfunction and neurodegeneration in humans. The identification of shared clinical features and mitochondrial respiratory chain impairment provides valuable insights into the pathogenesis of this condition.

## MATERIALS AND METHODS

### Subjects

Patients were recruited at different hospitals and genetic testing laboratory worldwide, and were identified through GeneMatcher platform.^29^ Demographic and clinical data (biochemical analysis, neuroimaging acquisition and neurophysiological studies) were collected by each attending physician by retrospective review of clinical charts and registered on a standardized form. Brain Magnetic Resonance Images (MRI) were acquired as part of the routine clinical assessment in each case and five studies were systematically reviewed by an expert pediatric neuroradiologist (M.C.) to detail localization and characteristics of the lesions when present. Brain MRI reports were available for the rest of the seven cases.

### Ethical issues

This study was approved by the ethical committees of each institution and is in accordance with the Helsinki Declaration of 1964, as revised in 2001. The study was approved by the HSJD ethics committee with the reference n° PIC-97-16.s. Family 4 was enrolled in a research protocol approved by the Institutional Review Board at the Icahn School of Medicine at Mount Sinai, Family 5 was enrolled in the Solve-RD - solving the unsolved rare diseases program by the European Commission and Family 6 was part of the URDCat program. Adult participants and guardians of children provided written informed consent for participation.

### Laboratory methods

Biochemical analyses in blood, urine, and cerebrospinal fluid (CSF) were performed as part of routine diagnostic assessment at each center. In **P7**, the analysis of lactate, pyruvate, amino acids, and organic acids was conducted using automated spectrophotometric procedures, UPLC-MS/MS chromatography, and GC-MS/MS chromatography. The mitochondrial biomarkers FGF-21 and GDF-15 were analyzed in serum by ELISA as described by Dominguez-Gonzalez *et al*.^30^ Copper measurements in serum, CSF, and fibroblasts were performed by ICP-MS, as previously reported.^31^

### Genomics and bioinformatics

All patients were diagnosed by Next Generation Sequencing (NGS) at their institutions according to their established pipelines (more information is available on demand). Contact among different institutions were done through GeneMatcher platform^29^ and network of collaborators.

### mRNA expression analysis

mRNASeq was performed on **P7** fibroblasts. Quality control, library preparation, sequencing platform, and primary data analysis was done as previously described.^32^ Analysis of the aligned data was done using DROP (Detection of RNAseq Outliers Pipeline) in order to detect aberrantly expressed genes, altered splicing events, and monoallelic expression (MAE) of rare variants.^33–35^ The cohort of 303 fibroblasts from patients with Mendelian disorders^35^ was used as controls.

### In silico analysis

*In silico* modeling of the CTR1 protein has been performed using PyMol Molecular Graphics System (v.2.4.1, Schrödinger, LLC) with AlphaFold model AF-O15431-F1 fitted to model 6M98.^26^ The CTR1 protein sequence (O15431) has been used as a BLAST query and model organism sequences have been selected for comparison. Protein alignment has been performed using Clustal Omega (EMBL-EBI).

### Cell culture

Skin-derived fibroblasts of **P7** and controls were maintained in MEM (1 g/L glucose, 10% fetal calf serum and 1% penicillin–streptomycin) and grown to confluence in 75 cm^2^ flasks. When indicated, **P7** fibroblasts were treated with 50 μM or 100 μM copper histidinate (CuHis, Farmàcia Avel·lí Xalabarder Miramanda, Pujades, Barcelona, Spain) for 24 h or 72 h, respectively.

### Western Blot

Fibroblasts were homogenized in RIPA lysis buffer containing protease and phosphatase inhibitors (Bio Basic Inc.). Electroblotting was performed in 4-15% gradient precast gels TGX (Bio-Rad Laboratories, Hercules, CA, USA). Proteins were visualized by immunostaining with specific antibodies using Odyssey LICOR fluorescent system. The antibodies used in this study were: anti-SLC31A1 (ab129067, Abcam, Cambridge, UK) and anti-GAPDH (ab8245, Abcam, Cambridge, UK). Membrane images were quantified with ImageJ software (Bethesda, MD, USA).

### High-resolution respirometry

High-resolution respirometry was performed using polarographic oxygen sensors in a two-chamber Oxygraph-2k system at 37°C according to the manufacturer’s instructions (Oroboros Instruments). Manual titration of OXPHOS inhibitors (Oligomycin, Antimycin) and uncouplers (CCCP) was performed using Hamilton syringes (Hamilton Company) as previously described.^36^ Data were recorded and analyzed using the DatLab software v5.1.1.9 (Oroboros Instruments). Two biological replicates were performed for each experiment.

### Statistics and data analysis

Graphs and gene lists of proteins related to Cu metabolism or transport were analyzed on R Statistical Computing environment (https://www.r-project.org/). Statistical analysis of the biological assays were performed using the two-tailed Student’s *t*-test to compare the means of two independent groups of normally distributed data. Data were reported as the mean ± SD. *p*-values lower than 0.05 were considered statistically significant.

## Supporting information

Supplementary Clinical Information

Supplementary Clinical Information

## Data availability

The data that support the findings of this study are available on a reasonable request to the corresponding author and patient caregivers. The data are not publicly available due to privacy and ethical restrictions.

## ACKNOWLEDGEMENTS

We would like to thank the patients and their families for their collaboration. MU is a student of the Master of Genetics and Genomics at Universitat de Barcelona. We would like to thank “Biobanc de l’Hospital Infantil Sant Joan de Déu per a la Investigació” integrated in the Spanish Biobank Network of ISCIII for the sample and data procurement.

## SUPPLEMENTARY MATERIAL

Supplementary material is available online.

