## Supplementary Figures for "CLINICAL AND MOLECULAR CHARACTERISATION OF SLC31A1-RELATED NEURODEVELOPMENTAL DISORDER"

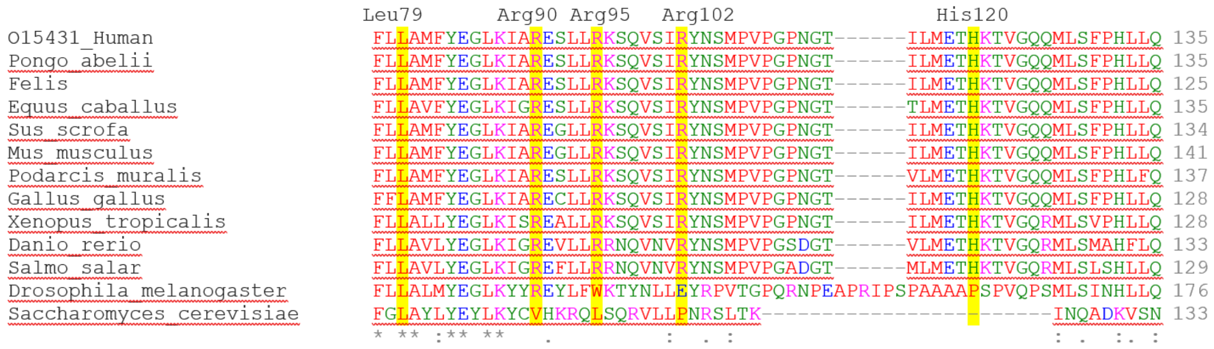


**Supplementary Figure 1. CTR1 protein multiple alignment.** hCTR1 and orthologues showing high conservation of the *SLC31A1* residues substituted in syndromic patients presented here and previously published in Batzios et al 2022[9] and Dame et al 2022.[10]

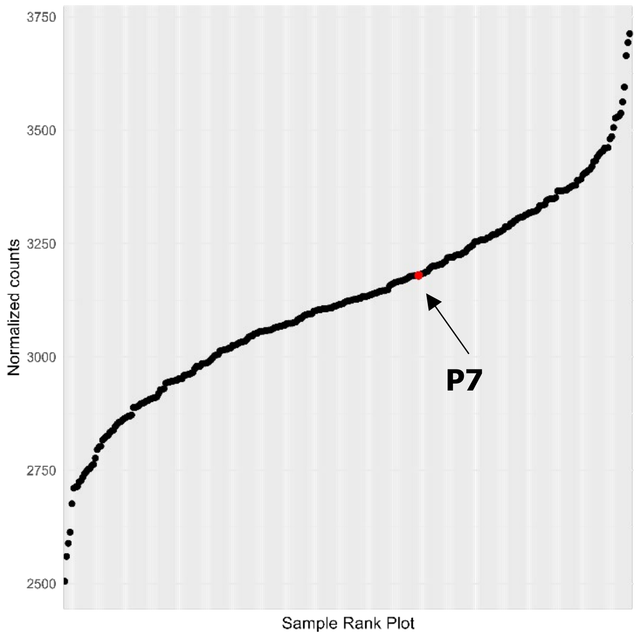


**Supplementary Figure 2. *SLC31A1* gene expression in P7.** Expression rank plot of *SLC31A1* gene. Red dot illustrates *SLC31A1* expression in **P7** fibroblasts, which had normal *SLC31A1* expression levels in comparison with the entire cohort of transcriptomes

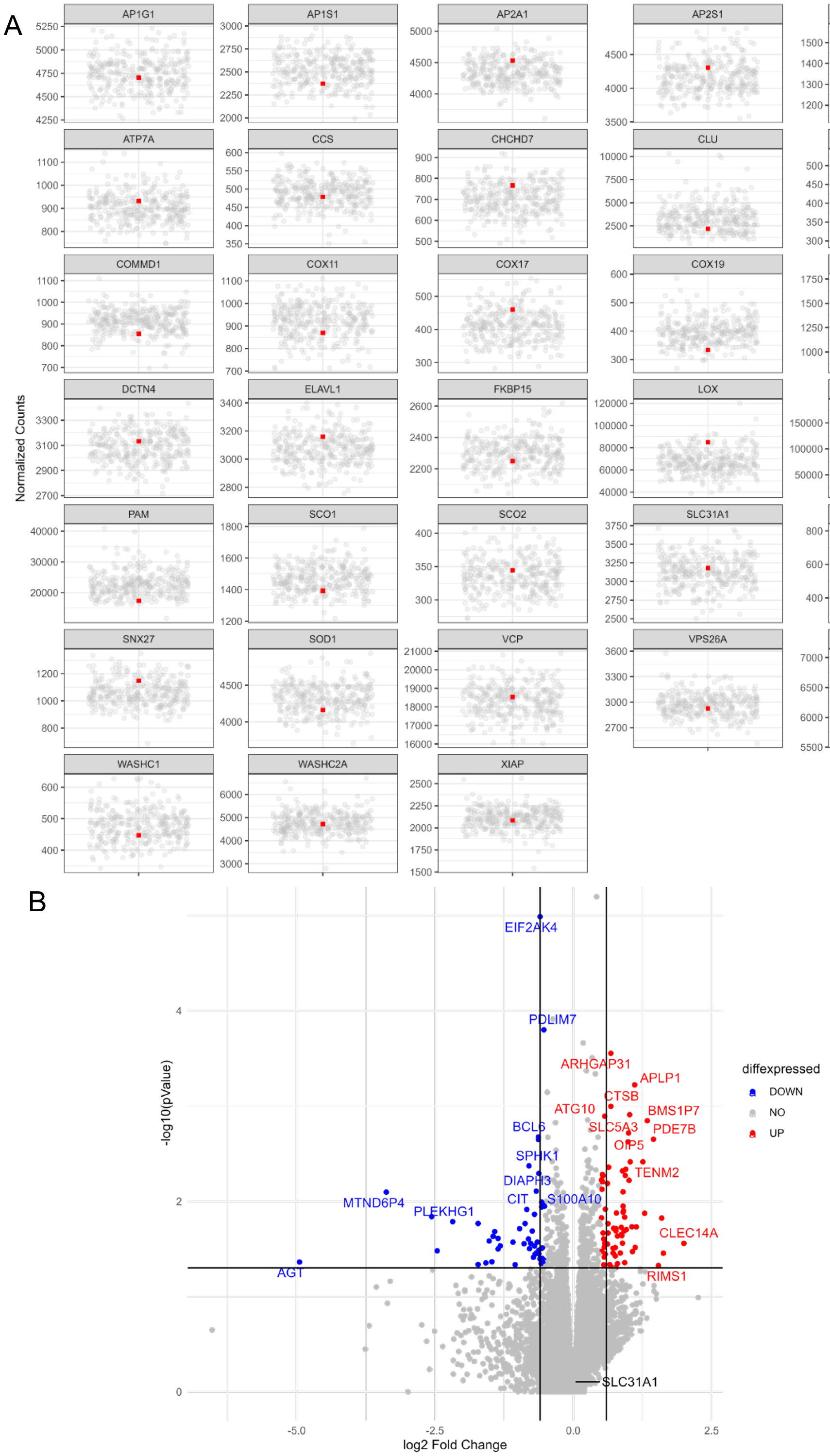


**Supplementary Figure 3. Cu-related genes expression levels.** (A) Gene expression levels of genes related to Cu metabolism or transport that have been seen altered in other copper-related pathologies. Red dot indicates **P7** normalized counts for the illustrated genes. (B) Volcano plot showing gene-level significance (−log10 p-Value) against log2 Fold Change. No aberrant outliers were prioritized in the sample.

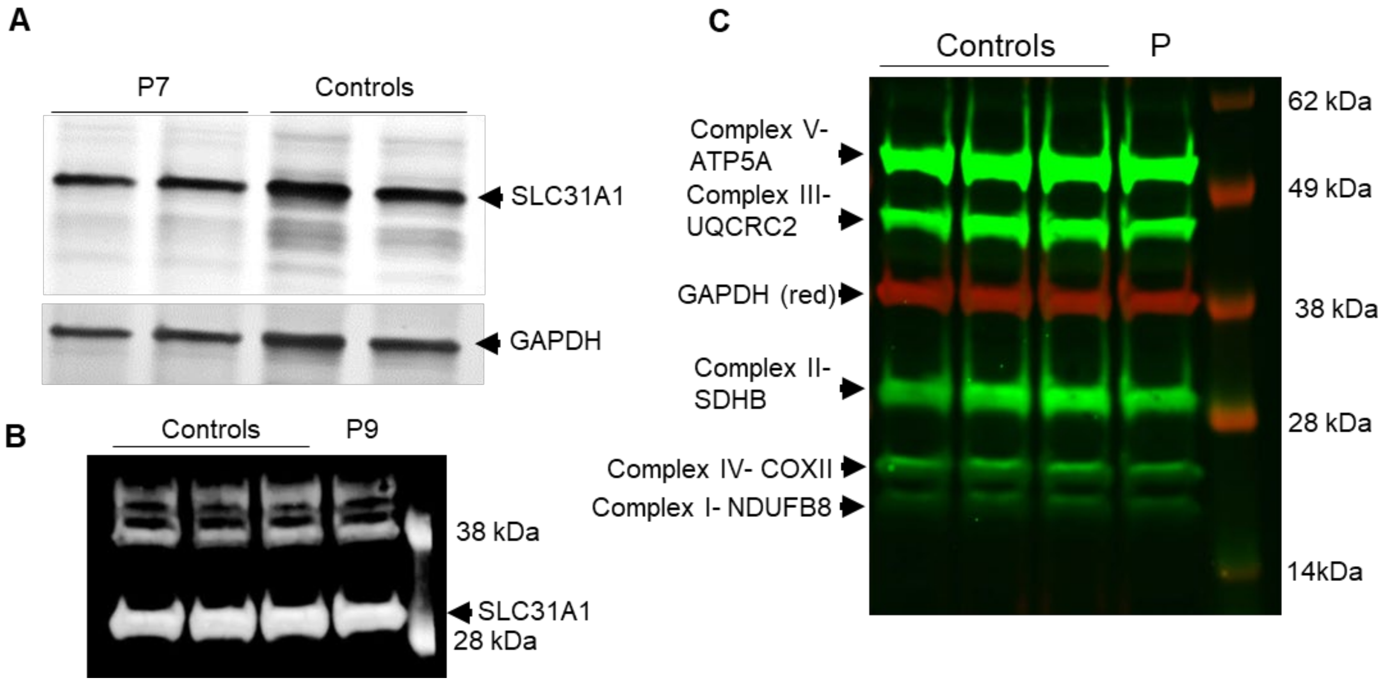


**Supplementary Figure 4. SLC31A1 and respiratory chain complex expression analysis by western blot.** SLC31A1 protein in **P7** fibroblasts **(A)** and **P9** lymphoblasts (**B**) compared with control samples. (**C**) Expression of respiratory chain complex, probed with anti-human oxphos complex antibody cocktail in **P9** lymphoblasts and controls.
