## Supplementary Clinical Information for "CLINICAL AND MOLECULAR CHARACTERISATION OF SLC31A1-RELATED NEURODEVELOPMENTAL DISORDER"

### Supplementary clinical description

To improve patient anonymization ages have been described in non-overlapping 6 months gaps (1-6, 7-12, 13-18, 19-24) and 2 years gaps for ages over 2 years (2-4, 5-7,8-10…)

#### Family 1 (P1)

Case P1 was the first child of consanguineous parents (Suppl. Fig. 1). The family history was notable for relatives being affected by epilepsy. She was born at term via vaginal delivery without complications. Anthropometric measures at birth were weight 3.350 kg (57th percentile), length 47 cm (5th percentile, -1.7 SD), and head circumference 34 cm (36th percentile). Seizures began at 1-6 months of age, followed by developmental regression and hypotonia, leading to a loss of social interaction, eye contact, head control, and vocalization. Initial seizures were characterized by upward gaze deviation for a few seconds. She developed intractable epilepsy with tonic and myoclonic seizures. Vigabatrin, topiramate, and primidone were ineffective in achieving seizure control. She subsequently exhibited severe global developmental delay and never achieved head control, sitting, or standing positions. Brain MRI at 0-6 months of age showed mild to moderate communicating hydrocephalus, enlargement of subarachnoid space, and diffuse increased T2 signal of cerebral white matter, suggesting delayed myelination. Follow-up brain MRI between 13-18 months of age showed cerebral and cerebellar atrophy, ventriculomegaly, and periventricular white matter hyperintensities. Laboratory investigations were unremarkable. Neurological exam at 5-7 years revealed no language development, severe axial hypotonia without head control, and muscle limb atrophy without deep tendon reflexes. Ophthalmological exam revealed bilateral blindness and optic disc pallor. Additionally, she had severe dysphagia and severe constipation. She died at 5-7 years.
Genetic testing revealed homozygosity for the c.283 C>T (p.Arg95Cys) variant in the *SLC31A1* gene.

#### Family 2 (P2)

The case was a 13-18 months male, born to a healthy consanguineous couple by vaginal delivery with forceps, after an uneventful pregnancy. His development appeared normal until 1-6 months of age, when it was observed that he could not support his head. He was subsequently admitted to the hospital for 2 weeks, during which all investigations were reported as normal, including a brain MRI (according to the family). A subsequent brain MRI revealed diffuse brain volume loss with bilateral cystic encephalomalacia within the frontal lobes, large chronic subdural hematomas at different stages, findings suggestive of axonal injury within the splenium of the corpus callosum, as well as hemorrhagic diffuse axonal injury within the left subinsular region and left frontal lobe. Hematological assessment was normal. On physical examination, the child had not acquired motor or social skills and exhibited respiratory distress. He had a flat occiput, depressed nasal bridge, and anteverted nostrils. He exhibited bilateral fisting, flexion deformities at the knees, head lag, axial hypotonia, and increased tone in the extremities. Additionally, he had severe oropharyngeal dysphagia, laryngomalacia, and gastroesophageal reflux. He experienced several episodes of severe respiratory infections requiring hospitalization, including intubation and assisted ventilation. During hospitalization, he presented with abnormal movements that could be compatible with seizures (not confirmed). Genetic testing revealed homozygosity for the *SLC31A1* c.284G>A (p.Arg95His) variant.

**Family 3 (P3)**

Case P3 was a male born to consanguineous parents (Suppl. Fig. 1) at 37 weeks of gestation by normal vaginal delivery after an uneventful pregnancy. Birth weight was 3,250 g (74th percentile), length 50 cm (78th percentile), and head circumference 36 cm (95th percentile, 1.68 SD). He exhibited motor developmental delay, managing to sit unsupported at 7-12 months but still unable to walk by 19-24 months. Focal seizures began at 19-24 years of age and progressed to epileptic encephalopathy refractory to treatment with phenobarbital. Developmental regression ensued, characterized by loss of previously acquired social and motor skills (smiling, eye contact, ability to hold his neck, and sitting) and hypotonia. The last neurological examination at 2-4 years revealed generalized hypotonia, tremors, and dysphagia; he did not exhibit social interactions (no social smile, no eye contact). Auditory brainstem response (ABR) evaluation around 13-18 months suggested retrocochlear hearing impairment, while ophthalmologic evaluation was normal. Neurological deterioration continued, and the patient died around the age of 2-4 years. He did not exhibit metabolic abnormalities, and 24-hour urine copper levels were within the normal range.
Genetic testing revealed homozygosity for the c.304C>T (p.Arg102Cys) *SLC31A1* variant.

**Family 4 (P4)**

The index case was a male patient born to healthy consanguineous parents. He presented with seizures starting at around 1-6 months of age, accompanied by a loss of head control and social communication. He exhibited hypotonia, global developmental delay, and failure to thrive. Additionally, Bilateral tonic-clonic seizure and progressive neurological deterioration were noted. He had subtle dysmorphic features consisting on downslanted palpebral fissures, an open mouth, micrognathia and low-set, slightly dysplastic ears. Brain MRI/MRS between 1-6 months of age showed bilateral symmetrical T2 and FLAIR hyperintensity targeting the basal ganglia, caudate, and lentiform nuclei, associated with hemorrhagic focus in the left caudate nucleus. Electrophysiological findings showed low amplitude of cortically recorded somatosensory evoked potentials. Laboratory investigations showed decreased circulating copper [21.4µg/dL; reference values (RV): 83-152] and ceruloplasmin (<1mg/dl; RV: 25-45) concentration, low urinary copper concentration (10.2µg/day; RV: 15-70), and high serum lactate (with a Lactate/Pyruvate ratio of 153:1). He had a similarly affected sibling presenting with pancytopenia and who died at 2-4 years of age with unexplained intracranial hemorrhage. He also has a healthy sibling.
The index patient is homozygous for two likely pathogenic variants: *ATP7B*(NM_000053.4):c.1646T>C; p.(Leu549Pro) and c.304C>T (p.Arg102Cys) in *SLC31A1*, both parents and the healthy brother were heterozygous for both variants (in *SLC31A1* and *ATP7B* genes). Unfortunately, segregation was not possible for the deceased sister.

**Family 5 (P5 and P6)**

**P5** was the third child of non-consanguineous parents. Two older siblings, aged 5-7 years, are healthy. Her perinatal history revealed an uneventful pregnancy, vaginal delivery at term with vacuum assistance, and a normal neonatal period. Birth parameters were within normal limits. Developmentally, she progressed normally until around 1-6 months, showing appropriate visual tracking and social smiling. However, at this time, developmental regression was observed, characterized by loss of head control and diminished visual tracking. She presented with infantile spasms, and an EEG showed disorganized and unstructured baseline activity, prompting the initiation of treatment with vigabatrin. Physical examination revealed retrognathia, inverted nipples, axial hypotonia with absent head control, and generalized chorea and dystonia. There was no eye contact or language development. Auditory brainstem responses were abnormal, consistent with neurosensory hearing loss. Oro-pharyngeal dysphagia for liquids was noted. Laboratory findings revealed increased CSF neopterin (59 nmol/L, RV: 12-30 nmol/L) along with elevated lactate levels in blood (3.20 mmol/L, RV: 0.77-2,44 mmol/L) and CSF (4.35 mmol/L, RV: 1-2,2 mmol/L ), suggesting a possible mitochondrial disorder, for which she received treatment with cofactors (biotin, thiamine, and carnitine). Other metabolic tests did not show significant abnormalities. Initial brain MRI around 7-12 months showed signal alterations in the thalamus and striatum, as well as in the corpus callosum, cerebellum, and midbrain, accompanied by cortico-subcortical volume loss and hypomyelination. T2-WI hyperintensity involving the pallidum and thalamus was observed, suggesting vigabatrin toxicity. Additionally, several superficial left temporal micro hemorrhagic foci were identified along with T2-WI hyperintensity of the cerebellar cortex corresponding to an old subarachnoid hemorrhage. As disease progressed, tonic seizures prompted treatment with zonisamide and levetiracetam. She had frequent hospitalizations due to bronchospasm crises. She experienced febrile status epilepticus during a respiratory infection episode. She passed away at 19-24 months.
Postmortem whole genome sequencing identified the variants *SLC31A1* c.304C>T (p.Arg102Cys) inherited from her father and c.363_364dupAA (p.Thr122LysfsTer8) of maternal origin.

Her younger sibling (**P6**) was born at term by vaginal delivery after an uncomplicated pregnancy. She had a normal perinatal period. Around 1-6 months of age, she exhibited infantile spasms, prompting treatment with vigabatrin, which was later switched to valproic acid. On physical examination, she was awake, with severe impairment of contact and comprehension. She occasionally babbled. She presented axial and limb hypotonia with choreiform movement in the hands. Laboratory findings showed increased lactate in blood (2.93 mmol/L, range 0.77-2,44 mmol/L) and cerebrospinal fluid (3.6 mmol/L, RV: 1-2,2 mmol/L). Brain MRI revealed T2 hyperintensity of the dentate nuclei of the cerebellum, diffusion restriction of the splenium of the corpus callosum suggesting vigabatrin toxicity. Enlarged perivascular spaces were also observed. Video EEG demonstrated multifocal epileptiform abnormalities (moderate incidence during wakefulness and high incidence during sleep) with diffuse distribution (predominance in frontal and parietal areas) and a slight tendency to spread. As mitochondrial disease was suspected, mitochondrial cofactors were initiated. She passed away at 7-12 months of age from respiratory insufficiency during a bronchospasm crisis. She exhibited initial symptoms at the same age as her sister, but her condition deteriorated more quickly, resulting in an earlier death.

Sanger sequencing of the variants found in her sister confirmed the diagnosis.

**Family 6 (P7)**

P7 was the first child of a non-consanguineous, healthy family (Suppl. Fig. 1). He was born at term after an uneventful pregnancy. Anthropometric measures at birth were weight 3,490 g (59th percentile), length 53 cm (94th percentile), and head circumference of 36.5 cm (84th percentile). Severe delayed psychomotor development was noted shortly after birth, with no social smile, eye contact, or head control. Around 1-6 months of age, he began to experience epileptic spasms, with an EEG showing slow baseline activity with isolated spikes in the left parieto-temporal lobe. He was treated with levetiracetam (50 mg/kg/day), topiramate (3 mg/kg/day), and ACTH (the latter was administered for two months, with some transient improvement in psychomotor development and intermittent social smiling), but seizures persisted. Around 1-6 months of age, he was admitted to the Pediatric ICU due to acute respiratory failure caused by a *P. carinii* respiratory infection, requiring non-invasive mechanical ventilation. In the following months, the patient experienced polymorphic seizures, including oral automatisms accompanied by ictal vomiting, tonic, tonic-clonic, myoclonic, and partial clonic seizures. EEG analysis revealed a slow and disorganized background with multifocal epileptiform discharges. His seizures were refractory to treatment with oxcarbazepine, carbamazepine, rufinamide (which appeared to worsen his seizures), levetiracetam, phenobarbital, lamotrigine, topiramate, vigabatrin, zonisamide, adrenocorticotropic hormone, and a ketogenic diet. In addition, bilateral neurosensory hearing impairment was detected. Routine laboratory tests, including a complete blood count, blood chemistry, and thyroid function, were mostly normal. Metabolic workup showed persistently elevated plasmatic lactate (4.7-6.4 mmol/L; RV: 1.1-2.2), pyruvate (0.195 mmol/L; RV: 0.03-0.1), alanine (675 µmol/L; RV: 167-439 µmol/L). Urine organic acids revealed increased excretion of Krebs cycle intermediates (malate and fumarate) and 2-OH-isobutyric acid. CSF lactate, pyruvate, and alanine were also elevated (3.98 mmol/L; RV: 1.0-2.22, 0.175 mmol/L; RV: 0.064-0.136, and 66 µmol/L; RV: 12-35, respectively). Additional mitochondrial biomarkers, such as growth differentiation factor 15 (GDF-15) and fibroblast growth factor-21 (FGF-21), were mildly elevated (625 pg/ml; RV: 200-540 and 534 pg/ml; RV: 10-300, respectively). Serum copper (983 µg/L, RV: 620–1544 µg/L) and ceruloplasmin (229 mg/dL, RV: 200–360) were within normal ranges. Under suspicion of a mitochondrial disease, measurement of mitochondrial respiratory chain enzyme activities in frozen muscle identified a mild deficit of complex II, complex II + III, and a significant deficiency in complex IV (cytochrome C oxidase) activity (21.7, control range: 88-180% activity/citrate synthase Units). Muscle biopsy showed marked variation in myofiber size with atrophy and multiple vacuoles corresponding to small round lipid droplets within most of the fibers. Brain MRI at 7-12 months of age showed T2-WI hyperintense signal abnormalities in the basal ganglia and diffuse brain atrophy, more prominent in the bilateral frontotemporal lobes, with secondary ventricular dilation and bilateral frontal and right occipital subdural hygroma. Hyperintense signal abnormalities were observed in the bilateral brainstem, with severe temporal cortical-subcortical atrophy and significant enlargement of the subarachnoid space of the anterior temporal fossa, along with thinning of the corpus callosum. Follow-up MRI 13-18 months of age showed severe frontotemporal cortical-subcortical atrophy with significant enlargement of the subarachnoid space of the bihemispheric convexity associated with frontoparietal subdural hygromas. Compensatory enlargement of the ventricular system and prominent thinning of the corpus callosum were also observed. Diffuse signal abnormalities, with supratentorial white matter and basal ganglia hyperintensity on T2-weighted imaging, were detected. Spectroscopy of the left lentiform nucleus showed reduced NAA with a slight increase in

choline and traces of lactate. At 2-4 years of age brain MRI showed significant cerebral atrophy and findings of probable diffuse sclerosis in both cerebral hemispheres, especially in the parieto-occipital and central regions, in the basal ganglia, brainstem, and dentate nuclei of both cerebellar hemispheres. On examination at this age, the child was unresponsive while awake, without spontaneous movements or vocalization, and progressive microcephaly was observed (<2SD). Profound truncal hypotonia and weakness persisted, with severe motor delay: the child never acquired head support and required a gastrostomy tube for nutritional support. He died at 2-4 years of age from cardiorespiratory arrest during epileptic decompensation.
Genetic testing by whole exome sequencing revealed the homozygous variant c.360C>G (p.His120Gln) in the *SLC31A1* gene.

**Family 7 (P8, P9 and P10)**

The proband (**P9**, II.2 in Family 7, Suppl. Fig 1) is a 5-7 years old male born before 35 weeks of gestation to non-consanguineous parents. Vaginal delivery was induced due to maternal pre-eclampsia. Birth weight was 2,543 g (70th percentile). He began experiencing seizures around the age of 1-6 months, characterized by tonic arm movements and eye deviation. Brain MRI at this age revealed chronic subdural hematomas, severe supratentorial volume loss with enlargement of the ventricular system. At this point, he was able to roll over; however, by 13-18 months of age, he exhibited severe hypotonia with no head control and no eye contact. Microcephaly and dysmorphic facial features were observed, including bitemporal narrowing, a mildly flat nasal bridge with an upturned nose, strabismus, and large earlobes. He required a tracheostomy and gastrostomy tube for respiratory and nutritional support, respectively. He has no known cardiac, gastrointestinal, genitourinary, or endocrine issues. Follow-up MRI at that moment showed progression of the supratentorial volume loss, with ventriculomegaly, diffuse prominence of the subarachnoid spaces, and increased T2 signal in the lenticular nuclei. The proband was analyzed by WES, identifying the variant c.360C>G (p.His120Gln) in the SLC31A1 gene in homozygosity.
His sibling (**P10**, II.3 in Family 7, Suppl. Fig 1) had a similar clinical course, including epileptic encephalopathy with refractory infantile spasms, severe global neurodevelopmental delay, and hypotonia. Additionally, he had cortical visual impairment and a patent foramen ovale. At 1-6 months, neuroimaging revealed supratentorial volume loss with extensive signal intensity on T2-WI, most pronounced in the frontal and anterior temporal lobes, involving the subcortical U fibers, along with bilateral moderate-sized subdural collections likely attributed to rapid parenchymal volume loss. EEG demonstrated epileptiform activity with intermittent spikes at F3, F4, and CZ. A13-18 months he was non-verbal and unable to walk. Sanger sequencing confirmed the presence of the same variant found in the proband.
Their sibling (**P8**, II.1 in Family 7, Suppl. Fig 1) had a history of severe global developmental delay, never achieving any developmental milestones, along with epilepsy and hydrocephalus that required a ventriculoperitoneal shunt. He passed away at 2-4 years of age. Postmortem Sanger sequencing confirmed homozygosity for the c.360C>G variant in the SLC31A1 gene. The variant was absent in the healthy sibling.

**Family 8 (P11)**

Case P11 (II.3 in Family 8, Suppl. Fig. 1) is 11-13 years old, residing in a long-term care facility due to complex medical needs for over a decade. Although the family denies consanguinity, SNP array results suggest a potentially inbred background. She was born following an uncomplicated delivery with a birth weight of 3,900 g (91st percentile), and Apgar scores of 7 and 9, being discharged home after 2 days. Within the first few months of life, she developed intractable seizures (both generalized and myoclonic), accompanied by profound hypotonia, severe global developmental delay, and microcephaly. She has been ventilator-dependent due to chronic respiratory failure since 7-12 months of age. Metabolic workup included tests for plasma lactate, amino acids, very long-chain fatty acids (VLCFA), transferrin electrophoresis, and alpha-N-acetyl-D-glucosaminidase activity. These tests were selected to help diagnose disorders associated with genes located in the regions of homozygosity identified, but results were normal. Urine organic acids were normal, but cerebrospinal fluid (CSF) lactate was mildly elevated. Brain MRI between 7-12 months of age showed severely diminished parenchymal volume with ex-vacuo enlargement of the ventricular system and subarachnoid spaces, a thin corpus callosum, and abnormal T2 hyperintense signals throughout the basal ganglia, thalami, cerebral white matter, upper left parietal lobe, and medial cerebellar hemispheres, with scattered susceptibility foci in many of these areas. Homozygosity for variant c.360C>G (p.His120Gln) in the SLC31A1 gene was determined by WES.

Her sibling (II.4 in Family 8, Suppl. Fig. 1), who was more severely affected, presented with early-onset epileptic encephalopathy beginning at 1-6 months of live, leading to multiple hospitalizations due to recurrent status epilepticus. Initial metabolic workup showed elevated lactate at 3.7 mmol/L (reference values: 1.1-2.2 mmol/L), but subsequent tests indicated normal lactate levels on two occasions. TORCH studies yielded negative results, and CSF studies revealed normal glucose, protein, and amino acid levels. Plasma amino acids, acylcarnitine profile, and urine organic acids were also normal. Brain MRI at 1-6 months of age showed diffuse cerebral atrophy with abnormal signals in the thalami. EEG revealed a generalized slow background. She passed away between 7-12 months of age due to status epilepticus. Autopsy reported moderate microcephaly with a cone-shaped cranium, polygyria, and neuronal loss in deep gray matter, with a brain weight of 375 g. Neuropathology revealed severe bilateral neuronal loss and gliosis in the caudate nucleus, putamen, and thalami. Genetic testing to confirm an SLC31A1-related disorder was unsuccessful, as only very fragmented DNA was obtained from formalin-fixed paraffin-embedded (FFPE) tissues from the autopsy material.

**Family 9 (P12)**

The index case was born to healthy, consanguineous parents. She had a sibling of 8-10 years who had a language delay. She was delivered at full term via cesarean section. At birth, her length, weight, and head circumference (HC) were around the 50th percentile. However, at follow-up, she was small for her age with an HC of 43.5 cm (<1 percentile; -2.8 SD). She presented with failure to thrive, severe developmental delay, absent language, microcephaly, and hypotonia in both upper and lower limbs, with head lag. She could move her extremities and hold objects but was unable to sit or stand. She could recognize her mother and interacted by smiling. Around 7-12 months of age, she was diagnosed with a Wilms tumor, which was treated with surgery and chemotherapy. Metabolic follow-up showed normal results for plasma amino acids, creatine phosphokinase, urine organic acids, carnitine, and homocysteine. The first newborn screening was remarkable for a C14:1 level of 0.5 μmol/L (cut-off = 0.4 μmol/L), but the repeat screening showed normal levels. Brain MRI at 13-18 months revealed diffuse abnormalities in the white matter, corpus callosum, and thalami, with foci of diffusion restriction. Follow-up MRI at 2-4 years showed persistent diffuse abnormalities in these regions, with interval improvement and residual restricted diffusion in the left external capsule, along with brain atrophy. There were no intracranial metastases. Ophthalmologic examination revealed healthy discs and flat retinae but bilateral cataracts. Electroretinography (ERG) was normal, while flash visual evoked response (VER) studies were abnormal. EEG findings included diffuse high-amplitude disorganized delta slowing, frequent bursts of high-voltage spikes, and wave discharges with inter-burst periods of severely attenuated delta activity. The patient expired at 2-4 years of age.

Whole exome sequencing (WES) analysis revealed that the patient is homozygous for the c.358C>T (p.His120Tyr) variant in the SLC31A1 gene. Additionally, she is homozygous for the NM_000018.4:c.1246G>A (p.Ala416Thr) variant in the very long-chain acyl-CoA dehydrogenase gene (*ACADVL* * 609575), which is associated with (VLCAD) deficiency (ACADVLD; MIM # 201475). This *ACADVL* variant is classified as pathogenic based on the ACMG criteria (PS1, PM1, PM2, PP3, PP5). This particular variant has been previously described as associated with mild or late-onset (adult form) of ACADVL deficiency, and functional analysis revealed that it maintains moderate activity.[1-4] Consequently, the ACADVL variant has not been considered responsible for the main clinical presentation of the patient. However, we cannot rule out that it may have influenced her clinical outcome to some extent.

**Family 10 (P13)**

The index case was born to consanguineous healthy parents. Prenatal history is remarkable for intrauterine growth restriction (IUGR) with a weight, length and head circumference of 2.5 kg (3rd percentile), 45 cm (2 percentile; -2.0 SD) and 33 cm (11th percentile) respectively. The affected individual presented with failure to thrive and profound developmental delay; she did not attain unsupported sitting, walking, or speech milestones. She showed irritability and disturbed sleep without other behavioral alterations as well as feeding difficulties. Physical examination was remarkable for dysarthria, hypertonia, muscle weakness, spasticity, ataxia, dystonia and limb contractures, thoracic kyphosis, hearing impairment and eye movement abnormalities. Ophthalmologic findings included optic disc pallor. She began experiencing myoclonic seizures followed by tonic-clonic seizures at 1-6 months of age, which have been unresponsive to treatment with Levetiracetam, Carbamazepine, and Valproic acid. The seizures lasted between 1 to 5 minutes and did not occur in clusters. Developmental delay was observed prior to seizure onset, with a noticeable regression of milestones beginning at that time. MRI at that moment showed cerebral atrophy and communicating hydrocephalus. At 5-7 years she was alive and presents with low wight (<3rd percentile) and short stature (<3rd percentile) without microcephaly (25th percentile). She presents with severe global developmental delay, speech delay, dystonia, hypertonia and is able to walk with support.

Genetic testing revealed homozygosity for the likely pathogenic variant c.559G>T (p.Glu187*) in the *SLC31A1* gene.
